# Humoral immunogenicity of the seasonal influenza vaccine before and after CAR-T-cell therapy

**DOI:** 10.1101/2021.05.10.21256634

**Authors:** Carla S. Walti, Andrea N. Loes, Kiel Shuey, Elizabeth M. Krantz, Jim Boonyaratanakornkit, Jacob Keane-Candib, Tillie Loeffelholz, Caitlin R. Wolf, Justin J. Taylor, Rebecca A. Gardner, Damian J. Green, Andrew J. Cowan, David G. Maloney, Cameron J. Turtle, Steven A. Pergam, Helen Y. Chu, Jesse D. Bloom, Joshua A. Hill

## Abstract

Recipients of chimeric antigen receptor-modified T (CAR-T) cell therapies for B-cell malignancies are immunocompromised and at risk for serious infections. Vaccine immunogenicity is unknown in this population. We conducted a prospective observational study of the humoral immunogenicity of 2019-2020 inactivated influenza vaccines (IIV) in children and adults immediately prior to (n=7) or 13-57 months after (n=15) CD19-, CD20-, or BCMA-targeted CAR-T-cell therapy, as well as controls (n=8). Individuals post-CAR-T-cell therapy were in remission. We tested for antibodies to 4 vaccine strains at baseline and ≥1 time point after IIV using neutralization and hemagglutination inhibition assays. An antibody response was defined as a ≥4-fold titer increase from baseline at the first post-vaccine time point. Baseline A(H1N1) titers in the CAR-T cohorts were significantly lower compared to controls. Antibody responses to ≥1 vaccine strain occurred in 2 (29%) individuals before CAR-T-cell therapy; one individual maintained a response for >3 months post-CAR-T-cell therapy. Antibody responses to ≥1 vaccine strain occurred in 6 (40%) individuals vaccinated after CAR-T-cell therapy. An additional 2 (29%) and 6 (40%) individuals had ≥2-fold increases (at any time) in the pre- and post-CAR-T cohorts, respectively. There were no identified clinical or immunologic predictors of antibody responses. Neither severe hypogammaglobulinemia nor B-cell aplasia precluded antibody responses. These data support consideration for vaccination before and after CAR-T-cell therapy for influenza and other relevant pathogens such as SARS-CoV-2, irrespective of hypogammaglobulinemia or B-cell aplasia. Larger studies are needed to determine correlates of vaccine immunogenicity and durability in CAR-T-cell therapy recipients.

**Key Points:**

- Influenza vaccination was immunogenic pre- and post-CAR-T-cell therapy, despite hypogammaglobulinemia and B-cell aplasia.
- Vaccination with inactivated vaccines can be considered before CAR-T-cell therapy and in individuals with remission after therapy.

## INTRODUCTION

The development and approval of chimeric antigen receptor-modified T (CAR-T) cell therapies for lymphoma, leukemia, and multiple myeloma (MM) is leading to wider-scale use in children and adults.^1–3^ These individuals are profoundly immunocompromised from their underlying malignancy and prior anti-tumor treatments, in addition to CAR-T-cell therapy related factors including lymphodepleting chemotherapy and cytokine release syndrome (CRS).^4^ Severe and often persistent cytopenias occur in part due to “on-target/off-tumor” depletion of non-malignant B-lineage cells expressing the CAR-T-cell targets.^1–7^

Strategies to prevent infections after CAR-T-cell therapy are not well established. Many patients are treated with prophylactic immunoglobulin replacement therapy (IGRT), which consists of pooled immunoglobulin G (IgG) isolated from blood from over 1,000 donors.^8^ However, there is limited evidence to support the efficacy of prophylactic IGRT in this context, and IGRT is primarily beneficial for prevention of only serious bacterial infections.^9^ Vaccination is a potentially more cost-effective and durable approach to infection prevention for some pathogens, but there are no published data regarding vaccine immunogenicity in CAR-T-cell therapy recipients. Vaccine immunogenicity, while often lower in immunocompromised patients compared to healthy individuals, is often nonetheless beneficial. For example, influenza vaccination in immunocompromised patients may be associated with lower rates of influenza infection and lower respiratory tract disease, a reduction in hospitalization, and lower mortality.^10,11^

Understanding vaccine immunogenicity in the context of CAR-T cell therapy is critically important to guide infection prevention strategies. These data are particularly relevant given the availability of vaccines for severe acute respiratory syndrome coronavirus 2 (SARS-CoV-2). Vaccination before treatment, as is preferred in solid organ transplant recipients,^12^ may be particularly important as the B-cell depletion that results from CAR-T-cell therapy may further abrogate immunogenicity. Vaccination starting 3-6 months after CAR-T-cell therapy is advocated by current guidelines,^13,14^ but the recommendation is extrapolated from other patient populations and treatments.^15–20^

Respiratory tract infections, particularly with viruses, are the most common infectious complication after CAR-T-cell therapy, and influenza has been reported as a cause of death.^20–23^ Among patients with cancer and hematopoietic cell transplant (HCT) recipients, influenza causes substantial morbidity and mortality with death occurring in 11% to 33% of affected individuals.^11^ Thus, there is an urgent need to understand the specific utility of influenza vaccination prior to and after CAR-T cell therapy, and to inform the broader question of vaccine immunogenicity in this patient population.

We report the results of a prospective observational study of the humoral immunogenicity of the seasonal inactivated influenza vaccine (IIV) among CD19-, CD20-, and BCMA-targeted CAR-T-cell therapy recipients vaccinated before or after CAR-T-cell therapy compared to controls.

## METHODS

### Study design and participants

We enrolled 3 distinct cohorts in the fall and winter of 2019-2020. We approached all children and adults planning to receive an IIV (1) prior to CD19-, CD20- or BCMA-CAR-T-cell therapy (pre-CAR-T cohort; IIV administered after leukapheresis and ≥2 weeks prior to CAR-T-cell therapy per institutional practice) at Fred Hutchinson Cancer Center (Fred Hutch) or Seattle Children’s Hospital (SCH), and (2) in remission after CAR-T-cell therapy without initiating new anti-neoplastic therapies (post-CAR-T cohort). The third cohort included Fred Hutch employees between 18 and 64 years of age who received an IIV through occupational health, were not immunocompromised, and volunteered to participate in the study (control cohort). Individuals who received IGRT within 2 months prior to enrollment were excluded. This study was approved by the Fred Hutch Institutional Review Board; all participants provided informed consent in accordance with the Declaration of Helsinki.

### Inactivated influenza vaccines

Individuals received a commercially available trivalent or quadrivalent 2019-2020 Northern Hemisphere IIV. Vaccines in the CAR-T cohorts are detailed in **Table 1**. All controls received a quadrivalent IIV (Flucelvax, Seqirus). The World Health Organization (WHO) recommended strains were: A/Brisbane/02/2018 (H1N1)pdm09-like virus, A/Kansas/14/2017 (H3N2)-like virus, and B/Colorado/06/2017-like virus (B/Victoria/2/87 lineage) for the trivalent IIV, with the addition of a B/Phuket/3073/2013-like virus (B/Yamagata/16/88 lineage) for the quadrivalent IIV.^24^

**Table 1.**
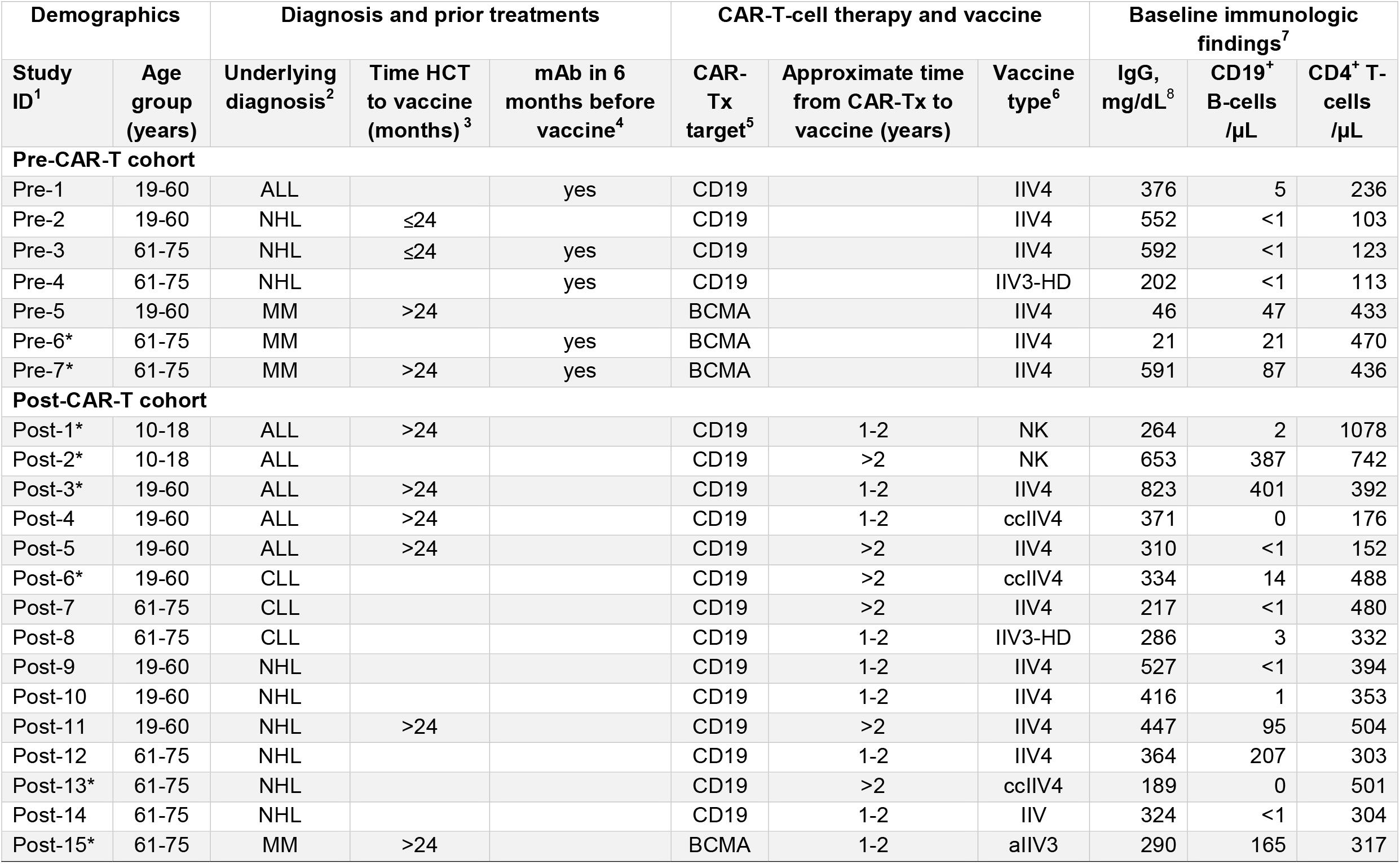
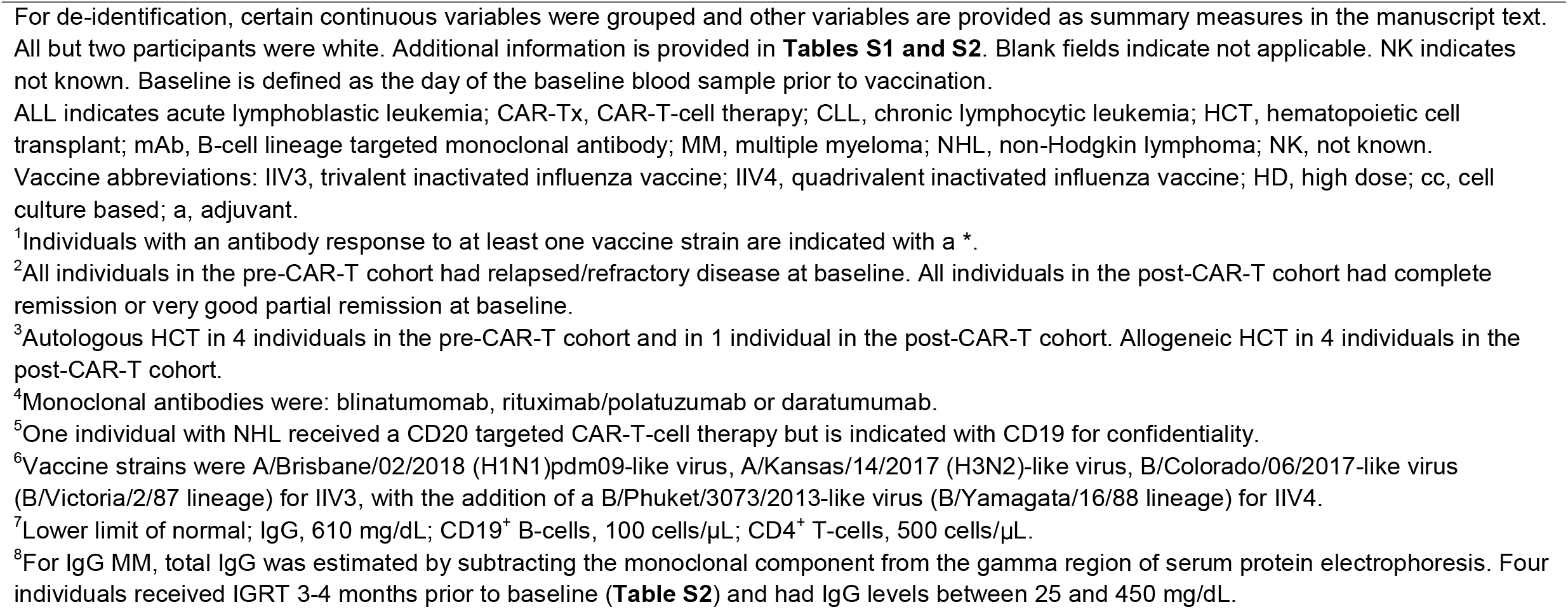
Baseline clinical characteristics and immunologic findings of the pre- and post-CAR-T-cell therapy cohorts.

### Data and blood collection

For the CAR-T-cell cohorts, data were abstracted from medical records. IGRT within 4 months (≥4 half-lives of circulating IgG) before any study sample collection was documented because of the potential for influencing measured antibodies.^25,26^ For the control cohort, date of birth, sex, and information about influenza vaccination in the prior year were collected.

In the pre-CAR-T cohort, blood samples were obtained before vaccination (baseline), before lymphodepleting chemotherapy, and approximately 30 and 90 days after CAR-T-cell therapy (**Figure 1**). In the post-CAR-T cohort, samples were collected at baseline and once approximately 30-90 days after vaccination. No samples were collected after relapse or start of new anti-tumor therapies. In the control cohort, samples were obtained at baseline and approximately 30, 60, and 90 days after vaccination. Serum and peripheral blood mononuclear cells (PBMCs) were isolated and stored (**Supplementary Methods**). Laboratory work was blinded to clinical characteristics.

**Figure 1.**
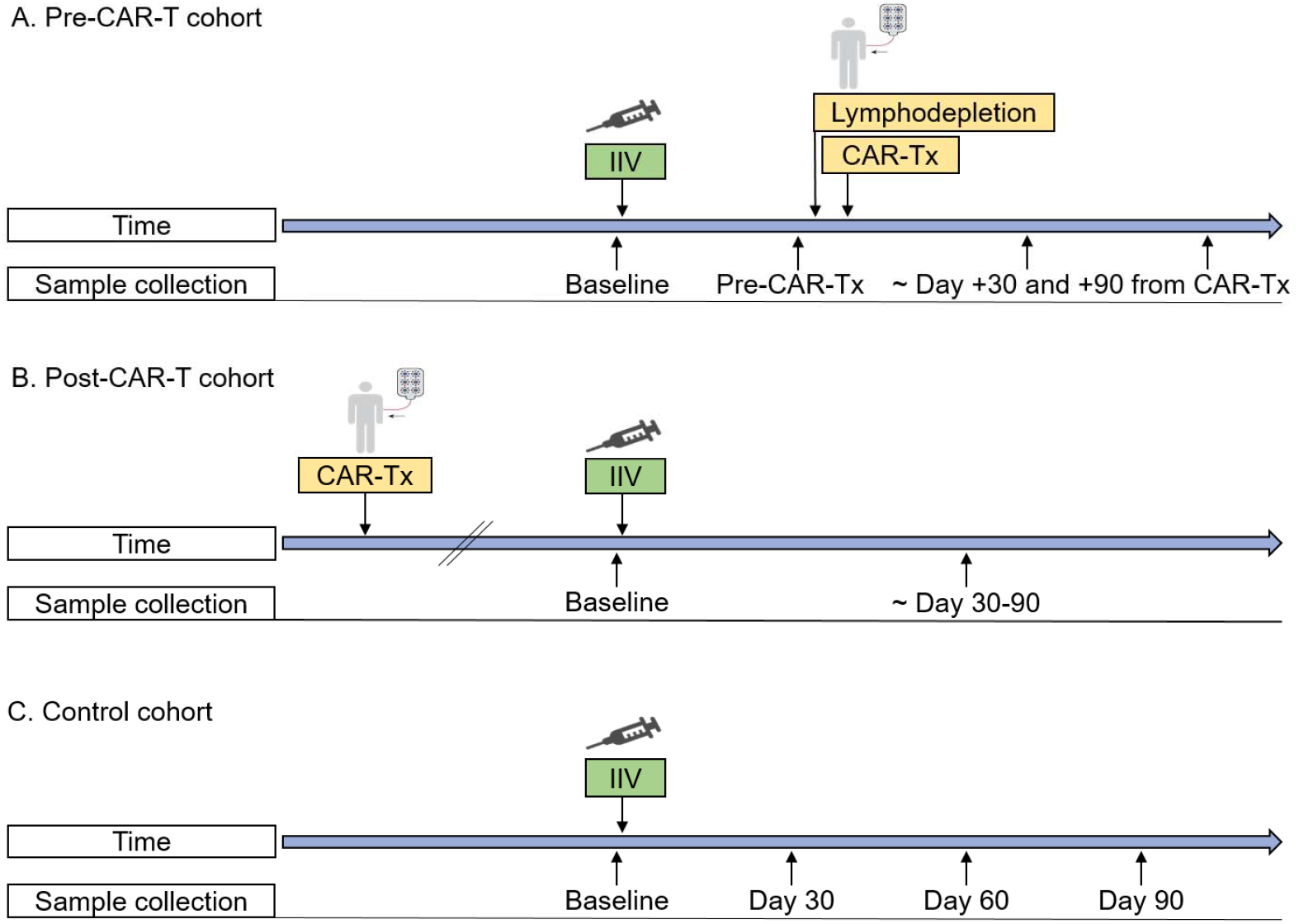
Inactivated influenza vaccine administration and sample collection timelines. Timelines demonstrating blood sample collection, inactivated influenza vaccine (IIV) administration, and CAR-T-cell therapy (CAR-Tx) for the (**A**) pre-CAR-T cohort (**B**), post-CAR-T cohort, and (**C**) control cohort. Lymphodepletion indicates lymphodepleting chemotherapy. In all cohorts, exact days of sample collection varied as detailed in the text and tables and depicted in figures.

### Laboratory testing

#### Hemagglutination inhibition (HAI) assay

Influenza hemagglutinin (HA) is the main target of neutralizing antibodies, and quantitation of HA-specific antibodies is the gold standard for measuring humoral immunity to influenza. We performed HAI assays on all serum samples and tested, in replicate serial 2-fold dilutions, for antibodies to all 4 vaccine strains as detailed in the **Supplementary Methods** and elsewhere.^27^ The highest dilution of serum that caused complete inhibition of hemagglutination was considered the titration end point. We reported the reciprocal of this dilution as the HAI titer. The lower and upper limits of detection (LOD) were 10 and 1280, respectively.

#### Neutralization assay

We also tested all serum samples with a fluorescent-based neutralization assay for antibodies against H1 of the A(H1N1) vaccine strain as previously described^28–30^ and detailed in the **Supplementary Methods**. This assay often correlates with the HAI assay and may be more sensitive. Additionally, it specifically measures neutralizing antibody activity, which might provide a better estimate of protection against infection.^31–33^ Two replicate dilution columns were used for each sample to calculate average infectivity. Neutralization was measured against virus containing the H1 sequence derived from the A/Brisbane/2/2018 H1N1pdm09 virus strain that carried a gene for the green fluorescent protein (GFP) in the PB1 segment. The assays were performed in MDCK-SIAT1-CMV-PB1 cells.^34^ Curves of fluorescence intensity were plotted and half maximal inhibitory concentrations (IC50s) were calculated using the *neutcurve* Python package. The IC50 is defined as the dilution of serum needed to inhibit infectivity of virus by 50% of its maximum infectivity as measured when no antibodies are present. We reported the reciprocal of IC50 as the neutralization titer. The lower and upper LOD ranged from 12.5-25 and 2680-5369, respectively.

#### Flow cytometry for B- and T-cells

We immunophenotyped B-cells and T-cells from PBMCs as detailed in the **Supplementary Methods**.

#### Total immunoglobulins

In addition to influenza-specific antibodies, we measured total serum IgG, IgM, and IgA using turbidometry (University of Washington Immunology Laboratory, Seattle, WA). In individuals with IgG MM, total functional IgG was estimated by subtracting the monoclonal component from the gamma region of serum protein electrophoresis.

### Outcomes

The primary outcome of interest for humoral immunogenicity from the IIV was an antibody response to the respective vaccine strains at the first post-vaccine time point. For the neutralization assay, we defined an antibody response as a ≥4-fold neutralization titer increase from baseline. For the HAI assay, we defined an antibody response as a titer of ≥40 if the baseline titer was <10 or a ≥4-fold rise from a baseline titer ≥10 (‘seroconversion’ as per the Food and Drug Administration [FDA]^35^). We separately reported the proportion of individuals with HAI antibody titers ≥40, a threshold often considered to correlate with seroprotection.^36^

### Analyses

We depicted absolute antibody titers at all time points in line and dot plots. A value of half of the lower LOD was assigned for values below the LOD. For each assay and vaccine strain, we compared baseline titers between cohorts using Kruskal-Wallis tests. If those tests were significant, Dunn’s test was conducted for pairwise comparisons using the Holm stepwise procedure to account for multiple comparisons. We calculated geometric mean titers (GMT) as summary measures. We described the proportion of individuals with an antibody response to each tested strain as defined above and with HAI titers ≥40. We also computed the proportion of individuals with an antibody response to ≥1 vaccine strain with Wilson 95% confidence intervals (CI). Post-vaccine HAI results for the B(Yamagata) strain were excluded for individuals without confirmed receipt of a quadrivalent vaccine. We used Spearman’s correlation to determine the correlation between the neutralization and the HAI assays for the H1N1 vaccine strain. We described differences in the primary outcomes by clinical and immunological characteristics. Two-tailed P values are reported. *P*<.05 was considered significant. All analyses were conducted using Stata (16.0).

### Data sharing

For original data, contact the corresponding author.

## RESULTS

### Baseline characteristics

We enrolled 30 children and adults: 7 in the pre-CAR-T cohort, 15 in the post-CAR-T cohort, and 8 in the control cohort. All received the IIV between September 2019 and March 2020. The most frequent vaccine type was the standard dose quadrivalent IIV. Clinical characteristics, baseline immunologic results, and vaccine information are detailed in **Table 1 and Tables S1 and S2**. The pre-CAR-T cohort included 7 adults with relapsed or refractory acute lymphoblastic leukemia (ALL; n=1), non-Hodgkin lymphoma (NHL, n=3), and MM (n=3). Four (57%) had a prior autologous HCT and 5 (71%) received a B-cell lineage targeted monoclonal antibody (mAb) therapy in the preceding 6 months. The post-CAR-T cohort included 2 adolescents and 13 adults who achieved a remission after receiving CAR-T-cell therapy a median of 21 months before IIV administration (range, 13-57 months). These individuals were treated for ALL (n=5), chronic lymphocytic leukemia (CLL; n=3), NHL (n=6), and MM (n=1). The majority of individuals in both CAR-T cohorts had hypogammaglobulinemia in addition to low absolute CD19^+^ B-cells and CD4^+^ T-cell counts. Controls were adults 25-62 years of age. The IIV was administered in the prior year to 13 (86%) individuals in the post-CAR-T cohort and all (100%) individuals in the control cohort; data were not reliably available for individuals in the pre-CAR-T cohort.

### Baseline influenza antibody titers

Baseline antibody titers in each cohort are depicted in **Figure 2** and summarized in **Table 2**. At baseline, neutralizing antibody titers to A(H1N1) were similar in the pre- and post-CAR-T-cell cohorts (GMT 26.5 vs. 45.4, *P*=.23) but were significantly higher in the control cohort (GMT 228.8; *P*=.01 compared to pre-CAR-T cohort, *P*=.02 compared to post-CAR-T cohort). These findings were similar using the HAI assay to A(H1N1), which demonstrated that antibodies at baseline were detectable in only 2 (29%) individuals in the pre- and 3 (20%) individuals in the post-CAR-T cohort compared to 7 (88%) in the control cohort. Correlation between the neutralization and HAI assay was high, but the neutralization assay was more sensitive (**Supplemental Results**). Baseline titers to A(H3N2) were low among all cohorts. Baseline titers to B(Victoria) or B(Yamagata) did not differ significantly between cohorts but tended to be slightly lower in the CAR-T-cell cohorts. Correspondingly, baseline HAI titers ≥40 to A(H1N1), B(Victoria), and B(Yamagata), but not to A(H3N2), were less frequent among CAR-T-cell therapy recipients than controls (**Table 2**).

**Figure 2.**
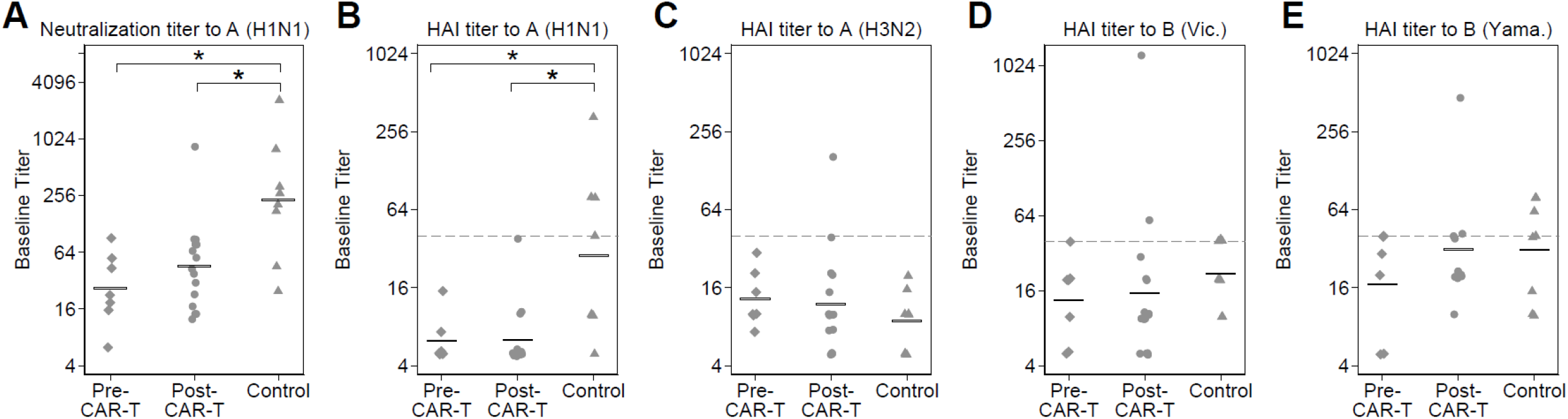
Baseline influenza antibody titers. Individual baseline titers are plotted by cohort for (**A**) the neutralization assay to A(H1N1), and the hemagglutination inhibition (HAI) assay to (**B**) A(H1N1), (**C**) A(H3N2), (**D**) B(Victoria), and (**E**) B(Yamagata). Data have been jittered to allow viewing of overlapping values. Horizontal bars represent geometric mean titers (GMTs). Points on or above the dashed horizontal lines represent baseline HAI titers ≥40. For both assays, titers to A(H1N1) were significantly lower in the CAR-T cohorts when compared to the control cohort as indicated with a * (neutralization assay: pre-CAR-T vs controls, *P*=.01; post-CAR-T vs controls, *P*=.02. HAI assay: pre-CAR-T vs controls, *P*=.009; post-CAR-T vs controls, *P*=.001; based on Dunn’s test with the Holm stepwise procedure for multiple comparisons). There were no significant differences between cohorts based on the HAI assay to A(H3N2), B(Victoria), or B(Yamagata) (Kruskal-Wallis, *P*=.46, *P*=.21 and *P*=.40, respectively), although in general, a higher proportion of individuals in the control cohort had HAI titers ≥40. GMTs and the proportion of individuals with an HAI titer ≥40 are detailed in **Table 2**.

**Table 2.**
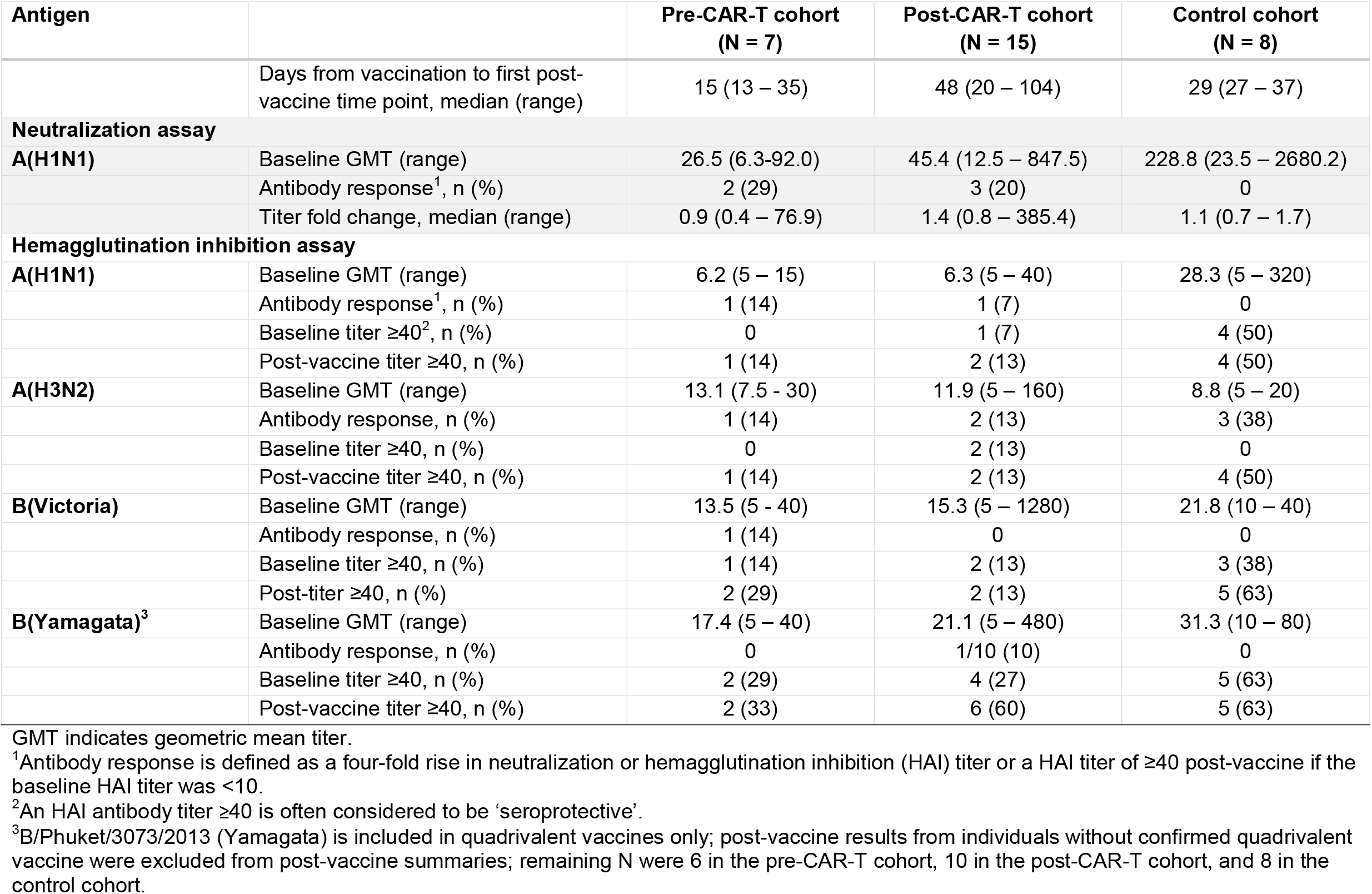
Antibody titers and antibody responses at baseline and at the first post-vaccine time point.

### IIV immunogenicity and kinetics of influenza antibody responses

#### Pre-CAR-T cohort

In the pre-CAR-T cohort (n=7), the IIV was administered a median of 35 days (range, 30-112) after the last dose of antineoplastic treatment, within a day after leukapheresis, a median of 0 days after baseline sample collection (range, 0-8), and 26 days (range, 14-50) before CAR-T-cell therapy. Two (29%) individuals received bridging antineoplastic therapy between leukapheresis and CAR-T-cell therapy. Five (70%) received treatment for immune related adverse events after CAR-T-cell therapy. By day 90 after CAR-T-cell therapy, 4 (57%) individuals achieved complete or very good partial responses of the underlying malignancy, 1 (14%) had persistent disease, and 2 (29%) died with progressive disease.

Plots of antibody titers over time for each strain are depicted in **Figure 3A**. At the first post-vaccine time point, a median of 15 days (range, 13-35) after IIV and before CAR-T-cell therapy, 2 (29%; 95% CI, 8%-64%) individuals demonstrated antibody responses to ≥1 vaccine strain (study ID, ‘pre-6’ and ‘pre-7’). Both had a response based on the neutralization assay to A(H1N1). These 2 individuals also had increased antibody titers based on the HAI assay to

**Figure 3.**
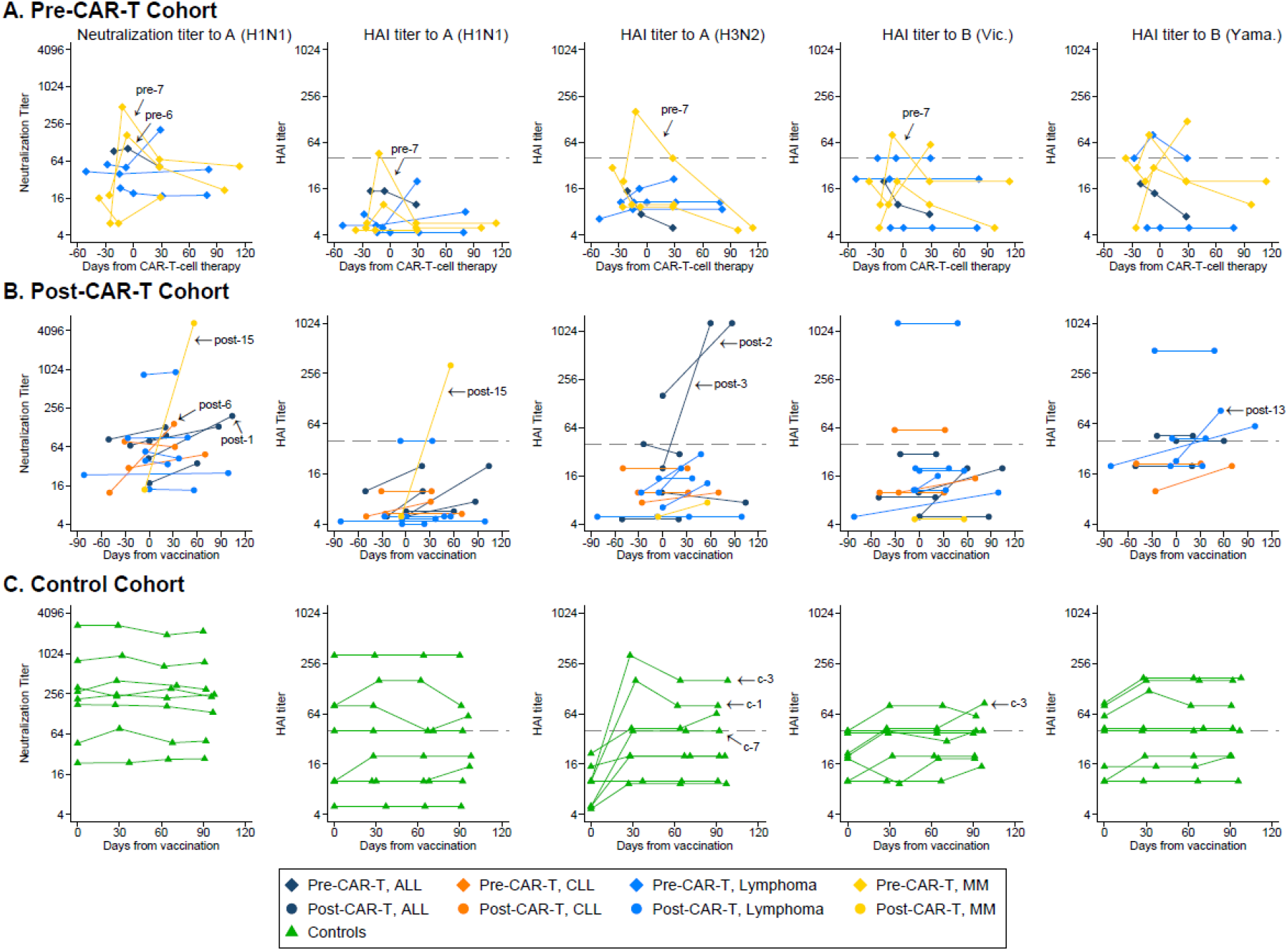
Kinetics of influenza antibody responses by individual. Line plots demonstrating neutralization titers to A(H1N1) and hemagglutination inhibition (HAI) titers to A(H1N1), A(H3N2), B(Victoria), and B(Yamagata) for (**A**) the pre-CAR-T cohort, (**B**) the post-CAR-T cohort, and (**C**) the control cohort. Each line connects results from one individual over time. Individuals with antibody responses per definition are indicated with an arrow and their study ID (**Table 1**). Symbols on or above the dashed horizontal line represent HAI titers ≥40. For the pre-CAR-T cohort, day 0 was set at the day of CAR-T-cell therapy, vaccines were administered between 0 and 8 days after baseline sample collection (median, 0), and time between vaccine and sample collection prior to CAR-T-cell therapy ranged from 13 to 35 days (median, 15). For the post-CAR-T cohort and the control cohort, day 0 was set at the day of vaccination. Individuals without confirmed receipt of a quadrivalent vaccine are excluded from the plots showing HAI titers to B (Yamagata).

A(H1N1) and to other strains, but only ‘pre-7’ met the HAI antibody response definition. After CAR-T-cell therapy, their titers decreased over time, but both still had a neutralization titer to A(H1N1) above baseline at ∼30 days after CAR-T-cell therapy, and ‘pre-7’ maintained a ≥4-fold increased titer for over 3 months. Both received immunosuppressive therapy for cytokine release syndrome and immune effector cell-associated neurotoxicity syndrome following CAR-T-cell therapy. Two other individuals (29%; ‘pre-3’ and ‘pre-5’) had ≥2-fold increases in antibody titers to two strains each at the first time point after CAR-T-cell therapy. Among non-responders, some had stable and some decreasing antibody titers over time. One individual (‘pre-4’) received IGRT after the first post-vaccine time point but had stable antibody titers at the next time point.

#### Post-CAR-T cohort

In the post-CAR-T cohort (n=15), the IIV was administered a median of 21 months (range, 13-57) after CAR-T-cell therapy (**Table 1**). All individuals were in ongoing remission. IgG was <400 mg/dL in 10 (66%) individuals; median CD19^+^ B-cell and CD4^+^ T-cell counts were 2.5 cells/μL and 392 cells/μL, respectively. The median time between the baseline sample collection and vaccination was 7 days (range, 0-82) and between vaccination and post-vaccine sample collection was 48 days (range, 20-104).

Plots of antibody titers over time for each strain are depicted in **Figure 3B**. Antibody responses to ≥1 vaccine strain occurred in 6 (40%; 95% CI, 20%-64%) individuals. Three (20%) individuals had an antibody response based on the neutralization assay to A(H1N1) and one of them also based on the HAI assay to A(H1N1). Three (20%) additional individuals had an antibody response to A(H3N2) or B(Yamagata). Six (40%) individuals did not meet response criteria but had a ≥2-fold increase to ≥1 strain each. Four (27%) individuals received IGRT within 62-95 days prior to the baseline sample, 3 of whom had subsequent IGRT within 23-71 days prior to the post-vaccine sample. Baseline titers in these individuals were similar to those who did not receive IGRT. One of three individuals who received IGRT between vaccination and post-vaccine time point had an antibody response which could have been affected by this measurement.

#### Control cohort

In the control cohort (n=8), the first post-vaccine time point was a median of 29 days from vaccination (range, 27-37 days). Plots of antibody titers over time for each strain are depicted in **Figure 3C**. All 3 (38%) individuals with an antibody response had increased titers for A(H3N2) only. Peak titers were observed at the first post-vaccine time point, and the responses were maintained through 90 days after vaccination. One of these individuals had a late ≥4-fold antibody titer increase to B(Victoria) at the 90-day time point. Three (38%) additional individuals had a ≥2-fold increase to ≥1 strain each.

#### Kinetics of influenza antibodies and GMTs for each cohort over time

Summary plots showing longitudinal antibody titers with GMTs for each cohort are depicted in **Figure 4**. This plot highlights a number of observations across the cohorts. Among both CAR-T cohorts, there was a modest increase in the GMT at the first post-vaccine time point. The pre-CAR-T cohort had a relatively rapid decrease in the GMT over time to a level below the baseline by the 90-day time point. Some individuals in the post-CAR-T cohort generated antibody titers as high or higher than the controls. The IIV for the 2019-2020 season had relatively low immunogenicity in the controls aside from strain A(H3N2), the strain to which no controls had a pre-vaccine HAI titer ≥40. Post-vaccine HAI titers ≥40 were more frequent in controls than in either CAR-T cohort.

**Figure 4.**
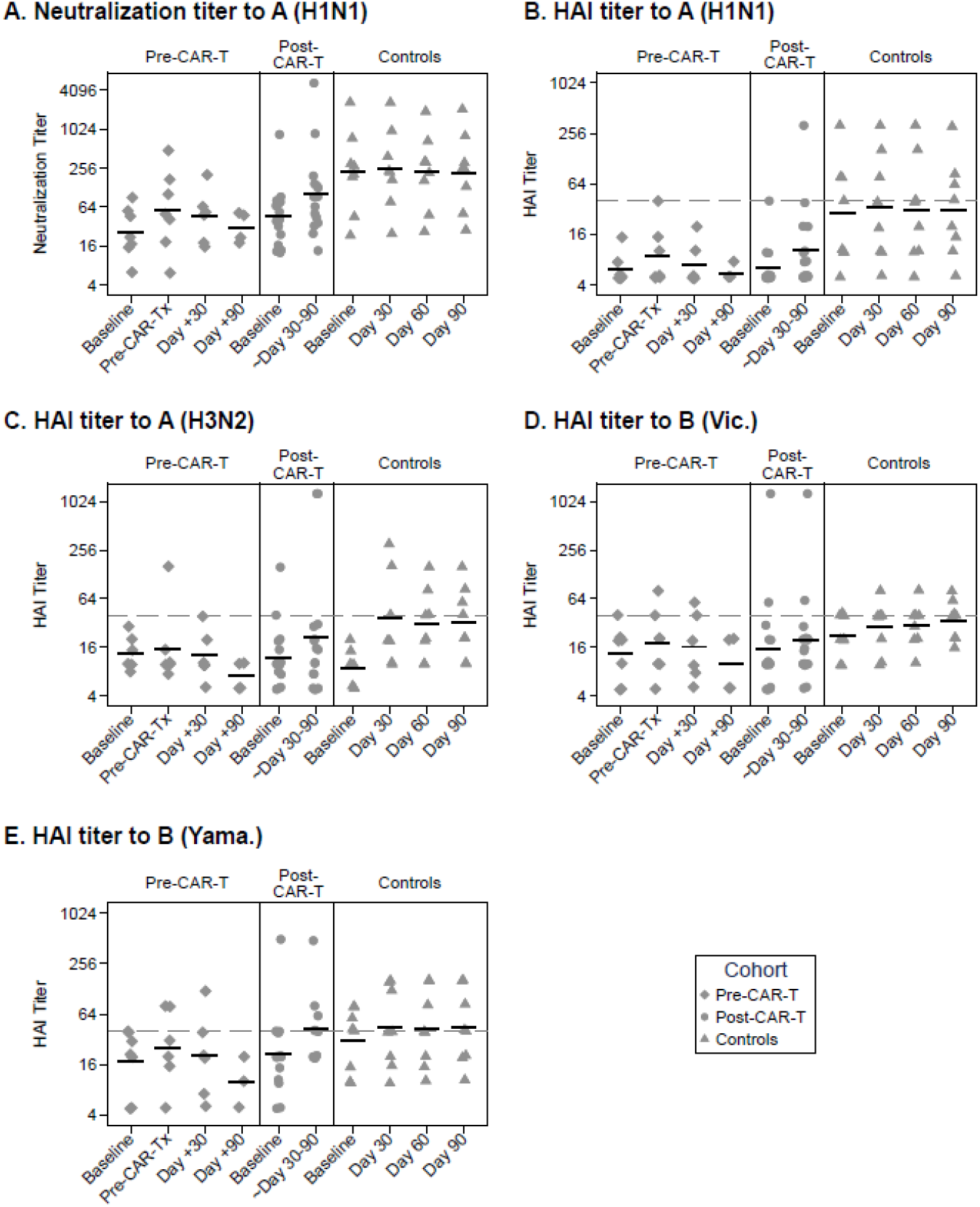
Summary of longitudinal influenza antibody kinetics and geometric mean titers for each cohort. Individual titer results are plotted per sample collection time points for the pre-CAR-T, post-CAR-T, and control cohorts (from left to right in each panel). (**A**) Neutralization titers to A(H1N1) and (**B**) hemagglutination inhibition (HAI) titers to A(H1N1), (**C**) A(H3N2), (**D**) B(Victoria), and (**E**) B(Yamagata) are shown. Data have been jittered to allow viewing of overlapping values. Horizontal bars represent geometric mean titers (GMT). Symbols on or above the dashed horizontal line represent HAI titers ≥40.

### Correlates of IIV immunogenicity

To explore possible correlates of IIV immunogenicity, we determined the clinical and immunologic characteristics of individuals who did and did not generate antibody responses. We depicted the fold-changes in neutralizing and HAI antibody titers for the pre- and post-CAR-T cohort in **Figure 5**, stratified for key baseline clinical and immunologic characteristics. Overall, there were no apparent correlates of antibody responses in either CAR-T-cell therapy cohort with evidence of immunogenicity across most categories of clinical and immunologic characteristics. Although only individuals with MM in the pre-CAR-T cohort had responses, and neither of the 2 individuals with CD4^+^ T-cell counts <200 cells/μL had responses, these observations are limited by small numbers. Importantly, in the post-CAR-T cohort, antibody responses were observed in individuals with very low peripheral CD19^+^ B-cells (including one individual with no detectable CD19^+^ B-cells at baseline) and individuals with severe hypogammaglobulinemia (IgG <400 mg/dL). All or most individuals with an antibody response had IgA and IgM levels below the lower limit of normal, respectively (**Table S1**). Additional clinical characteristics, baseline immunologic results, and IIV information of responders and non-responders are described in the **Supplement, Table 1, Table S1 and S2**.

**Figure 5.**
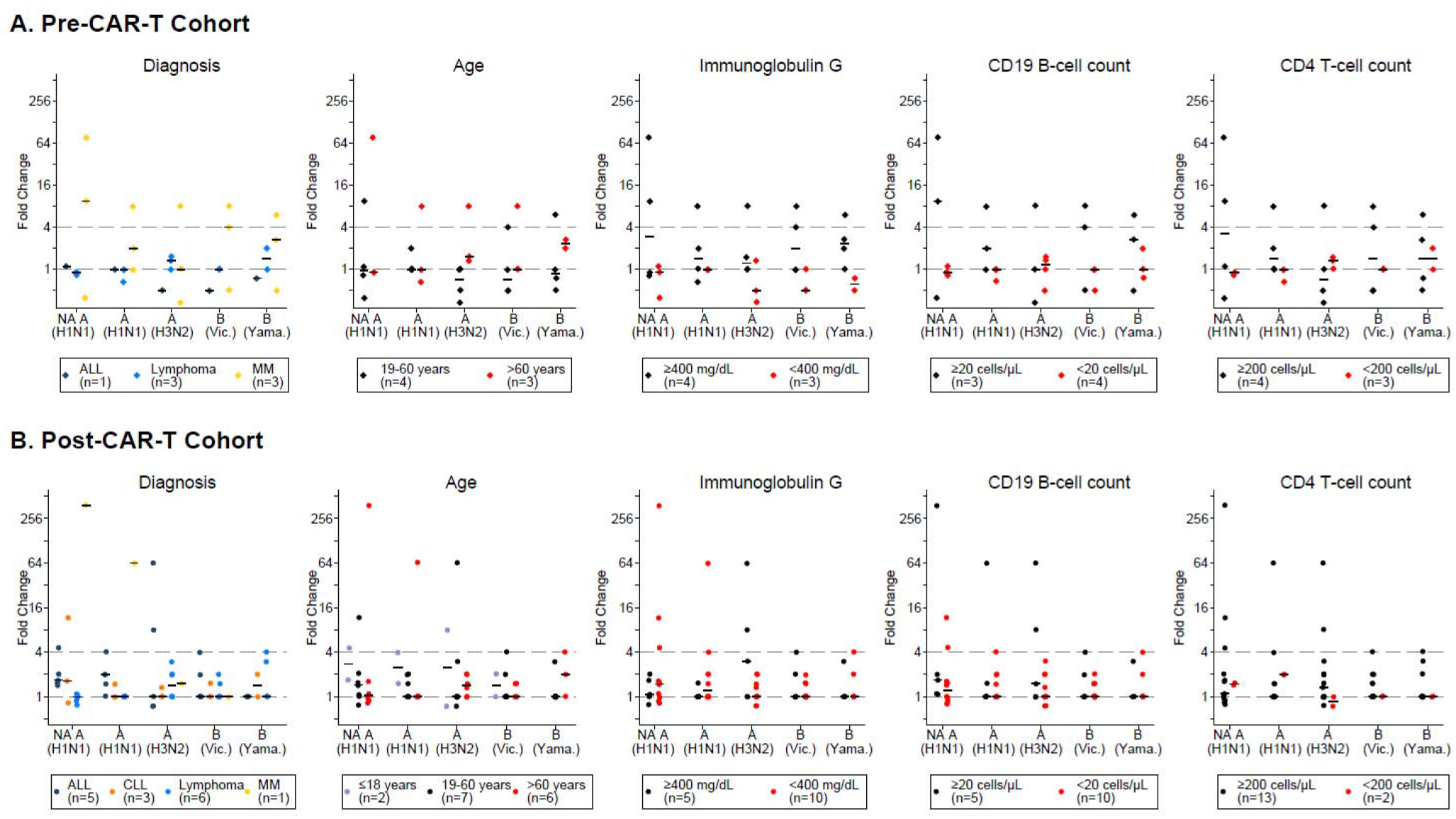
Antibody titer fold-changes by baseline clinical characteristics and immunologic findings. Antibody titer fold-changes from baseline to the first post-vaccine time point are depicted for each vaccine strain in (**A**) the pre-CAR-T cohort and (**B**) the post-CAR-T cohort. Each panel is stratified by a baseline characteristic (diagnosis, age, immunoglobulin G, CD19^+^ B-cell count, and CD4^+^ T-cell count). Characteristics are specified in the figure legends and indicated with different symbol colors. NA indicates neutralization assay; other results are based on hemagglutination inhibition (HAI) assays to A(H1N1), A(H3N2), B(Victoria), and B(Yamagata). A fold-change of 1 (lower dashed horizontal line) indicates no change in antibody titer from baseline. Horizontal bars represent median fold-changes. Symbols on or above the upper dashed horizontal line represent ≥4 fold-changes.

## DISCUSSION

Development of humoral immunity in response to vaccination plays an important role in protection against infection and severe disease^37^ as recently underscored by the SARS-CoV-2 pandemic.^38,39^ CAR-T-cell therapy recipients are highly immunocompromised prior to and for months following therapy, rendering them high-risk for infections.^4,7,23,40,41^ Vaccination may be an effective strategy to prevent the acquisition and severity of infections,^13^ but there are no reported data about vaccine immunogenicity, or predictors of responses to vaccines, in this patient population. Nonetheless, certain factors, such as hypogammaglobulinemia or low B-cell counts, are often considered when deciding upon the utility of vaccination. In this study of the IIV administered either shortly before CAR-T-cell therapy or in prior CAR-T-cell therapy recipients in remission, we demonstrated that 60-80% of individuals in both cohorts developed robust or partial antibody increases to ≥1 vaccine strain despite substantial humoral and cellular immunodeficiency. These findings support consideration for administration of relevant vaccines before CAR-T-cell therapy and for (re)vaccination, as indicated, of individuals in long-term remission, irrespective of serum IgG level and total B-cell count.

Immunity to influenza at baseline, prior to vaccination, reflects an individual’s history of prior exposure to vaccines and natural infection and can exhibit (cross-)reactivity to current vaccine strains. The 2019/2020 H1N1 vaccine strain only differed by a few amino acids compared to the 2018/2019 formulation.^24^ This may explain the high baseline antibody titers to A(H1N1) in the controls, all of whom were vaccinated in the prior year.^42–45^ In contrast, we demonstrated a high proportion of undetectable baseline titers to A(H1N1) in individuals pre-CAR-T-cell therapy, which may be due to lack of vaccination in the prior year, poor responses to prior vaccination, or loss of pre-existing immunity related to their malignancy and its treatment.^15,16^ Among individuals in remission after CAR-T-cell therapy, baseline titers to A(H1N1) were also significantly lower than in controls despite a similarly high frequency of prior year vaccination, suggesting either poor responses and/or rapid waning due to inability to establish long-lived antibody-secreting plasma cells.^46,47^ Baseline antibody titers to the A(H3N2) vaccine strain were low among all cohorts, likely due to a new A(H3N2) strain in the 2019/2020 vaccine formulation.^24^ Both 2019/2020 influenza B vaccine strains were unchanged from the previous year formulation and there was a trend towards lower baseline titers in the CAR-T-cell cohorts. Overall, a higher proportion of controls had HAI titers ≥40 for most strains, demonstrating that the CAR-T-cell cohorts may have higher risk for morbidity from influenza infection.^37,48^

After receiving the IIV, 29% of the pre-CAR-T cohort, 40% of the post-CAR-T cohort, and 38% of the controls had ≥4-fold increases in antibody titers for ≥1 vaccine strain, most of whom also developed a post-vaccine HAI titer ≥40. Sixty to 80% of individuals in all groups had a ≥2-fold increase, and smaller increases in antibody titers may nonetheless be clinically relevant and provide some protection from infection or disease.^37,48^ The relatively limited responses to some strains could be reflective of the known phenomena that individuals with higher baseline titers and vaccination in the prior year have lower responses to subsequent vaccination against the same strain, as evidenced by the absent responses to A(H1N1) and the most pronounced responses to A(H3N2) in the controls.^42–45,49^ Among responders, peak titers generally occurred at the first post-vaccine time point. In the pre-CAR-T-cell cohort, this was prior to CAR-T-cell therapy, and we observed a relatively rapid antibody decay after CAR-T-cell therapy. Given that the 2 individuals with antibody responses received plasma cell targeted BCMA-CAR-T-cells for MM, this observation may be related to destruction of newly generated influenza-specific antibody-secreting plasma cells by the expanding CAR-T-cells.^50,51^ However, in both responders, antibody titers generally persisted above baseline for at least 30 days and up to 4 months after CAR-T-cell therapy, which may provide at least some immunity during the period of highest immunosuppression and infection risk.^4,23,37,40,41,48^ As patients are often hospitalized during this period, the finding might be particularly relevant for respiratory virus outbreaks with nosocomial transmission like influenza and SARS-CoV-2.^52,53^ Overall, these findings are consistent with observations of relatively impaired vaccine immunogenicity in individuals being treated for hematologic malignancies or who received a HCT, with influenza vaccine response rates between 0%-60%,^11,15,16,54–57^ but still indicate sufficient immunogenicity to support vaccination.

Our study cohorts had heterogeneity in clinical characteristics and CAR-T cell products, but there were no clear correlations between clinical or immunologic characteristics and antibody responses. Key observations included demonstration of vaccine immunogenicity in individuals with low peripheral CD19^+^ B-cell counts (<20 cells/µL) and serum IgG (<400mg/dL), as well as low IgA and IgM levels. Although some guidelines and clinical heuristics would suggest not vaccinating the majority of individuals in our CAR-T-cohorts, we nonetheless demonstrate clinically relevant immunogenicity of the IIV.^15,16^ The observations in this study were consistent with our hypotheses that antibody responses to vaccines before or after CAR-T-cell therapy are biologically plausible based on studies demonstrating responses post-rituximab, even without measurable peripheral blood B-cells.^58–60^ This could be due to the presence of B-cells below the limit of detection in blood and the possibility of persistence or recovery of B-cells in lymphoid tissue or the bone marrow. Whether responses originated from de novo naïve B-cells or boosted memory B-cells is unclear.

This study has several strengths. To the authors’ knowledge, it is the first study of vaccine immunogenicity prior to or post-CAR-T-cell therapy. We performed a prospective study using neutralization and gold-standard HAI assays of longitudinally collected samples to demonstrate IIV immunogenicity in a diverse cohort of CAR-T-cell therapy recipients, in addition to controls. Our data support consideration for administration of non-live vaccines before CAR-T-cell therapy for influenza, and by extrapolation, to other relevant pathogens in this clinical context (e.g., SARS-CoV-2 and pneumococcus). Additionally, vaccinations should be offered to patients in remission after CAR-T-cell therapy as previously suggested.^13^ The primary limitation is the relatively small sample size, but these data set the foundation for larger trials of both immunogenicity and efficacy. Other limitations include that IIV was at the discretion of clinical providers, and vaccine types and timing of sample collection were variable based on clinical follow up. It is possible that undocumented influenza infection in between blood draws confounded measurements, but this is unlikely. None of the vaccinations occurred within the first year after CAR-T-cell therapy, and additional data are needed to determine immunogenicity in these earlier time periods. Although HAI titers ≥40 generally correspond to a 50% reduction in the incidence of infection,^31^ this is not established in immunocompromised individuals. Cellular responses are another critical component of immunity to influenza and other infections;^61^ T-cell responses were not studied in this analysis but may demonstrate additional utility of IIV in this population with impaired B-cell immunity.

In summary, these data support consideration for vaccination for influenza and other pathogens, such as SARS-CoV-2, before and after CAR-T-cell therapy, irrespective of hypogammaglobulinemia or B-cell aplasia. Larger studies are needed to determine predictors of vaccine immunogenicity and durability in CAR-T-cell therapy recipients. Additional strategies to prevent infections, like vaccination of close contacts and standard precautions, should remain the backbone of infection prevention in these high-risk individuals.

## Supporting information

Strobe checklist

## Data Availability

For original data, contact the corresponding author.

## Acknowledgments

The authors thank the study participants, the participants’ local provider for assistance with sample collection and medical records, in addition to Naomi Wilcox (statistician, University of Washington, Seattle, WA), Cassandra Job (BS, Fred Hutchinson Cancer Research Center, Seattle, WA), Jessica Hirianto (undergraduate student, Fred Hutchinson Cancer Research Center, Seattle, WA), Joyce Maalouf (clinical research manager, Fred Hutchinson Cancer Research Center, Seattle, WA), Jessica Morris (data management analyst, Fred Hutchinson Cancer Research Center, Seattle, WA), and Atif Bhatti (BS, Fred Hutchinson Cancer Research Center, Seattle, WA) for assistance with sample processing and data management.

This work was supported by grants from the Swiss National Science Foundation (P2BSP3 188162 to C.S.W.), the National Institutes of Health National Cancer Institute (NIH/NCI; U01CA247548 to J.A.H and P01CA018029 to D.J.G and C.J.T), the NIH/NCI Cancer Center Support Grants (P30CA0087-48 and P30CA015704-44), the American Society for Transplantation and Cellular Therapy (to J.B.), a Washington Vaccine Alliance Pilot Grant (to S.A.P.), and Juno Therapeutics (a Bristol-Myers Squibb company). J.D.B. is an Investigator of the Howard Hughes Medical Institute. The content is solely the responsibility of the authors and does not necessarily represent the official views of the funding institutions.

## Contribution

J.A.H., C.S.W., and S.A.P. designed the study; C.S.W., J.K.-C., J.A.H., and T.L. collected the data; A.N.L. and J.D.B. performed the neutralization assay; K.S., C.R.W., and H.Y.C performed the HAI assay; J.B. and J.J.T. performed the flow cytometry analyses; C.S.W., E.M.K., A.N.L., K.S., J.B., and J.A.H analyzed the data; E.M.K, C.S.W., K.S., and J.A.H. created the figures; C.S.W., A.N.L., K.S., E.M.K., J.B., S.A.P., H.Y.C., J.D.B., and J.A.H. interpreted the data; C.S.W., E.M.K. and J.A.H. drafted the initial manuscript. All authors contributed to the writing and revision of the manuscript and approved the final version.

