## Supplemental Material for "Humoral immunogenicity of the seasonal influenza vaccine before and after CAR-T-cell therapy"

### SUPPLEMENTAL METHODS

#### Specimens

Serum was isolated from whole blood collected in clot activator red top vacutainers and stored at -80°C. Peripheral blood mononuclear cells (PBMCs) were isolated by Ficoll Histopaque centrifugation from whole blood collected in acid citrate dextrose vacutainers, washed twice with phosphate-buffered saline, resuspended in a mixture of 90% fetal bovine serum and 10% dimethyl sulfoxide (DMSO), cooled at a controlled rate and stored in liquid nitrogen.

#### Laboratory testing

##### Neutralization assay

For the fluorescent-based neutralization assay, we used previously described protocols.^1–3^ To generate the viruses, a co-culture of 293T-PB1 cells and MDCK-SIAT1-TMPRSS2-PB1 cells were transfected with reverse-genetics plasmids which encoded the viral genomic segments, including a modified segment which included green fluorescent protein(GFP) in place of PB1 (encoded in a pHH-PB1flank-GFP plasmid), and a protein expression plasmid encoding TMPRSS2. We used the H1 sequence derived from the A/Brisbane/02/2018 (H1N1) virus strain. Other viral genes (PB2, PA, NP, NS, M, and NA) were derived from the A/WSN/1933 strain (kindly provided by Robert Webster of St. Jude Children’s Research Hospital). The virus containing culture supernatants were clarified, aliquoted and frozen at -80°C. Prior to running neutralization assays, sera were treated with receptor-destroying enzyme (RDE) to prevent virus from binding to residual sialic acids present in the serum. To do this, a vial of lyophilized RDE II (Seiken, Cat No. 370013) was resuspended in 20 mL PBS, then serum was diluted 1:4 into RDE solution, incubated at 37°C for 2.5 hr, and then heat-inactivated by incubating at 55°C for 30 min. RDE-treated sera were diluted down the columns of 96-well plate in neutralization assay medium (Medium 199 supplemented with 0.01% heat-inactivated FBS, 0.3% BSA, 100 U of penicillin/ml, 100 μg of streptomycin/ml, 100 μg of calcium chloride/ml, and 25 mM HEPES). Average infectivity was calculated from measurements from two replicate dilution columns for each serum. Plates were incubated at 37˚C for 1 hour to allow virus-antibody binding. Then, 5 × 10^4^ MDCK-SIAT1-CMV-PB1 cells were added to each well. The cells enabled the virus to express GFP upon infection. Wells without added serum were used to measure maximal infectivity in the absence of neutralization. Wells without cells were used to measure background fluorescence in viral supernatants. After 16-20 hours incubation at 37˚C, GFP fluorescence intensity was measured using an excitation wavelength of 485 nm and an emission wavelength of 515 nm (12-nm slit widths). Percent of maximal infectivity was calculated by subtracting background fluorescence signal from all wells and dividing the signal from serum-containing wells by the signal from corresponding wells without serum. Average infectivity over duplicate measurements were calculated. Curves were plotted and IC50s were calculated using the *neutcurve* Python package (https://jbloomlab.github.io/neutcurve/, 0.3.1).

##### Hemagglutination inhibition (HAI) assay

HAI assays were performed to all four vaccine strains (FR-1665, FR-1666, FR-1667, FR-1669, International Reagent Resource, Manassas, VA). Serum samples were treated with RDE (VWR, Radnor, PE: MSPP370013) overnight at 37° and subsequently heated at 56°C for 30 minutes to remove nonspecific inhibitors. Serum was then adsorbed with red blood cells to remove nonspecific agglutinins. We prepared two replicate serial 2-fold dilutions (starting from 1:10) of 50µL of each treated sample on a 96-well microtiter plate, added 25µL standardized concentrations of vaccine antigen, and incubated at room temperature for 15 minutes. We added 50µL standardized turkey red blood cells to all wells and allowed to settle at room temperature for 30 minutes prior to result assessment. All runs included antisera and positive and negative controls.

##### Flow cytometry for B- and T-cells

B-cells and T-cells were quantified using a research flow cytometry panel. Peripheral blood mononuclear cells (PBMCs) were incubated for 30 minutes on ice with antibodies contained in 100 µL of FACS buffer that consisted of 1x DPBS containing 1% newborn calf serum (Life Technologies). Cells were then washed and analysed on a FACSymphony (BD Bioscience). The following antibodies were included for cell labelling: fixable viability dye (FV), anti-CD45 BV510 (HI30, BD), anti-CD3 BV605 (UCHT1, BioLegend), anti-CD4 Alexa Fluor 488 (OKT4, BioLegend), anti-CD8 APC-H7 (SK1, BD), anti-CD16 BV711 (3G8, BD), anti-CD14 BV711 (M0P-9, BD), anti-CD38 BUV661 (HIT2, BD), anti-IgD BUV737 (IA6-2, BD), anti-IgM PerCP- Cy5.5 (G20-127, BD), anti-CD20 BUV395 (2H7, BD), anti-CD19 BV421 (HIB19, BD), anti-CD27 PE-Cy7 (LG.7F9, Thermo Fisher), and anti-EGFR APC (cetuximab, R&D Systems). Naïve B-cells (CD19^+^CD27^-^CD38^-^IgD^+^) and switched memory B-cells (CD19^+^CD27^+^IgD^-^) were delineated within the lymphocyte population. Analyses were performed using FlowJo™ Software version 10.7.1 (Ashland, OR). Proportions from flow cytometry were multiplied with absolute lymphocyte counts from complete blood cell count results to calculate absolute B- and T-cell counts.

### SUPPLEMENTAL RESULTS

#### Correlation between the neutralization and the HAI assay

There was a good correlation between antibody titers for the neutralization assay and the HAI assay to A(H1N1) (baseline, *r*=0.82; first post-vaccine timepoint, *r*=0.92; fold-changes between baseline and the first post-vaccine timepoint, *r*=0.70). The neutralization assay was more sensitive with fewer titers below the limit of detection (**Figure 3**).

#### Correlates of the inactivated influenza vaccine immunogenicity

In the pre-CAR-T cohort, the two individuals with antibody responses had multiple myeloma, but a third individual with multiple myeloma did not demonstrate any responses. The individual who responded to several vaccine strains had the longest interval from the last prior antineoplastic treatment (>90 days). Neither of the two individuals with an antibody response received bridging antineoplastic therapy. Both individuals received a standard dose quadrivalent vaccine. They had among the highest baseline CD19^+^ absolute B-cell counts in this cohort although still at very low levels (<21 cells/μL and 87 cells/μL) (**Table 1**).

In the post-CAR-T cohort, antibody responses occurred across a range of clinical characteristics including underlying disease, age, sex (not shown), prior HCT, CD19- or BCMA-targeted CAR-T-cells, and time post-CAR-T-cell therapy (**Table 1**). Importantly, antibody responses were observed in an individual without detectable CD19^+^ B-cells and in individuals with IgG, IgA and IgM values below the lower limit of normal. All individuals with antibody responses had ≥200 CD4^+^ T-cells/µL, but only 2 individuals had values lower than this.

### SUPPLEMENTAL TABLES

#### **Supplemental Table 1. Additional baseline demographics and clinical characteristics** of the pre- and post-CAR-T-cell therapy cohorts

|  | CAR-T-cell therapy | Pre-CAR-T cohort | |
| --- | --- | --- | --- |
| Study ID^1^ | **CAR-Tx product^2^** | **Treatment for immune related adverse events^3^** | **Disease status 90d post CAR-Tx** |
| Pre-CAR-T cohort | | | |
| Pre-1 | Investigational | Yes | CR |
| Pre-2 | Investigational |  | Persistent |
| Pre-3 | Axicabtagene ciloleucel | Yes | Progressive |
| Pre-4 | Axicabtagene ciloleucel | Yes | CR |
| Pre-5 | Investigational |  | Progressive |
| Pre-6* | Investigational | Yes | VGPR |
| Pre-7* | Investigational | Yes | VGPR |
| Post-CAR-T cohort | | | |
| Post-1* | Investigational |  |  |
| Post-2* | Investigational |  |  |
| Post-3* | Investigational |  |  |
| Post-4 | Investigational |  |  |
| Post-5 | Investigational |  |  |
| Post-6* | Investigational |  |  |
| Post-7 | Investigational |  |  |
| Post-8 | Investigational |  |  |
| Post-9 | Axicabtagene ciloleucel |  |  |
| Post-10 | Axicabtagene ciloleucel |  |  |
| Post-11 | Investigational |  |  |
| Post-12 | Investigational |  |  |
| Post-13* | Investigational |  |  |
| Post-14 | Axicabtagene ciloleucel |  |  |
| Post-15* | Investigational |  |  |
| No data missing. Blank fields indicate not applicable. CAR-Tx indicates CAR-T-cell therapy; CR, complete response; VGPR, very good partial response.  ^1^Individuals with an antibody response to at least one vaccine strain are indicated with a *.  ^2^All individuals received a standard lymphodepleting regimen with cyclophosphamide and fludarabine. Co-stimulatory domains were CD28 for axicabtagene ciloleucel, CD28/4-1BB for a CD20 targeting CAR-T-cell product used in one individual only, and 4-1BB for the remaining investigational products. Study protocols were: NCT03103971, NCT03277729, NCT03502577, NCT03338972, NCT02028455, NCT01865617, NCT03105336, NCT02631044.  ^3^Includes cytokine release syndrome and immune effector cell-associated neurotoxicity syndrome. Treatments were corticosteroids with or without tocilizumab. | | | |

#### Supplemental Table 2. Baseline laboratory values and most recent IGRT administration for the pre- and post-CAR-T-cell therapy cohorts

| Study-ID^1^ | CD8^+^ T-cells/µL | % naïve of all CD19^+^ B-cells^2^ | % switched memory of all CD19^+^ B-cells^2^ | IgA, mg/dL^3^ | IgM, mg/dL^3^ | IGRT between 2 and 4 months prior to baseline^4^ | IGRT between vaccination and first post vaccine timepoint |
| --- | --- | --- | --- | --- | --- | --- | --- |
| Pre-CAR-T cohort | | | | | | | |
| Pre-1 | 224 |  |  | 38 | <10 |  |  |
| Pre-2 | 399 |  |  | 67 | 27 |  |  |
| Pre-3 | 616 |  |  | 74 | <10 |  |  |
| Pre-4 | 350 |  |  | 26 | 13 |  |  |
| Pre-5 | 217 | 29 | 4 | <2 | <10 |  |  |
| Pre-6* | 359 | 41 | 33 | 5 | <10 |  |  |
| Pre-7* | 801 | 52 | 8 | 21 | 24 |  |  |
| Post-CAR-T cohort | | | | | | | |
| Post-1* | 450 |  |  | <2 | <10 | yes | yes |
| Post-2* | 537 | 70 | 5 | 74 | 69 |  |  |
| Post-3* | 759 | 47 | 2 | 7 | 126 |  |  |
| Post-4 | 147 |  |  | 2 | 10 | yes | yes |
| Post-5 | 104 |  |  | 3 | 10 | yes | yes |
| Post-6* | 262 |  |  | 3 | 21 |  |  |
| Post-7 | 223 |  |  | 3 | 10 |  |  |
| Post-8 | 401 |  |  | 34 | 10 |  |  |
| Post-9 | 531 |  |  | 37 | 19 |  |  |
| Post-10 | 247 |  |  | 33 | 10 | yes |  |
| Post-11 | 346 | 64 | 2 | 2 | 34 |  |  |
| Post-12 | 181 | 79 | 1 | 53 | 18 |  |  |
| Post-13* | 138 |  |  | 14 | 10 |  |  |
| Post-14 | 167 |  |  | 37 | 10 |  |  |
| Post-15* | 189 | 63 | 2 | 59 | 223 |  |  |
| IGRT indicated IgG replacement therapy.  ^1^ Individuals with an antibody response to at least one vaccine strain are indicated with a *.  ^2^ No values are missing. Percentage of B-cell subpopulations are only displayed among individuals with ≥20 CD19^+^ B-cells/µL.  ^3^ Lower limits of normal; IgA, 84 mg/dL; IgM, 40 mg/dL.  ^4^IGRT within 4 months (≥4 half-lives of circulating IgG) before any of the study sample collections is displayed. IGRT within 2 months prior to baseline was an exclusion criteria. | | | | | | | |
